# Impact of the COVID-19 restrictions on the epidemiology of *Cryptosporidium* spp. in England and Wales, 2015-2021

**DOI:** 10.1101/2022.09.26.22280357

**Authors:** JP Adamson, R M. Chalmers, D Rh Thomas, K Elwin, G Robinson, A Barrasa

## Abstract

**Background:** In England and Wales, cryptosporidiosis cases peak in spring and autumn, usually associated with zoonotic and environmental exposures (*Cryptosporidium parvum*, spring/autumn) and with overseas travel and water-based activities (*Cryptosporidium hominis*, autumn). Restrictions to control the COVID-19 pandemic prevented social mixing and access to swimming pools and restaurants for many months. Foreign travel from the UK also reduced by 74% in 2020. However, these restrictions potentially increased environmental exposures as people sought alternative countryside activities locally. To inform and strengthen surveillance programmes, we investigated the impact of COVID-19 restrictions on the epidemiology of *C. hominis and C. parvum* cases.

**Methods:** *Cryptosporidium*-positive stools, with case demographic data, are referred routinely for genotyping to the national Cryptosporidium Reference Unit (CRU). Cases were extracted from the CRU database (01 January 2015 to 31 December 2021). We defined two periods for pre- and post-COVID-19 restrictions implementation corresponding to the first UK-wide lockdown on 23 March 2020: “pre-restrictions” between week 1, 2015 and week 12, 2020, and “post restrictions-implementation” between week 13, 2020 and week 52, 2021. We conducted an interrupted time-series analysis, assessing differences in *C. parvum* and *C. hominis* incidence, trends and periodicity between these periods using negative binomial regression with linear-splines and interactions.

**Results:** There were 21,304 cases between 01 January 2015 and 31 December 2021 (*C. parvum* = 12,246; *C. hominis* = 9,058). Post restrictions-implementation incidence of *C. hominis* dropped by 97.5% (95%CI: 95.4%-98.6%; p<0.001). The decreasing incidence-trend observed pre-restrictions (IRR=0.9976; 95%CI: 0.9969-0.9982; p<0.001) was not observed post restrictions-implementation (IRR=1.0081; 95%CI: 0.9978-1.0186; p=0.128) due to lack of cases. No periodicity change was observed post restrictions-implementation. Where recorded, 22% of *C. hominis* cases had travelled abroad. There was also a strong social gradient, with those who lived in deprived areas experiencing a higher proportion of cases. This gradient did not exist post restrictions-implementation, but the effect was exacerbated for the most deprived: 27.2% of cases from the most deprived decile compared to 12.7% in the pre-restrictions period. For *C. parvum*, post restrictions-implementation incidence fell by 49.0% (95%CI: 38.4%-58.3%; p<0.001). There was no pre-restrictions incidence-trend (IRR=1.0003; 95%CI: 0.9997-1.0009; p=0.322) but a slight increasing incidence-trend existed post restrictions-implementation (IRR=1.0071; 95%CI: 1.0038-1.0104; p<0.001). A periodicity change was observed for *C. parvum* post restrictions-implementation, peaking one week earlier in spring and two weeks later in autumn. Where recorded, 8% of *C. parvum* cases had travelled abroad. The social gradient observed for *C. parvum* was inverse to that for *C. hominis*, and was stable pre-restrictions and post restrictions-implementation.

**Conclusion:** *C. hominis* cases were almost entirely arrested post restrictions-implementation, reinforcing that foreign travel is a major driver of seeding infections. Increased hand-hygiene, reduced social mixing, limited access to swimming pools and limited foreign travel affected incidence of most gastrointestinal (GI) pathogens, including *Cryptosporidium*, in the same period. *C. parvum* incidence fell sharply but recovered throughout the post restrictions-implementation period, back to pre-restrictions levels by the end of 2021; this is consistent with relaxation of restrictions, reduced compliance and increased countryside use. The effect on our results of changes in health-seeking behaviours, healthcare access and diagnostic laboratory practices post restrictions-implementation is uncertain, but it is likely that access to GPs and specimen referral rate to CRU decreased. Future exceedance reporting for *C. hominis* should exclude the post restrictions-implementation period but retain it for *C. parvum* (except the first six weeks post restrictions-implementation where the incidence fell sharply). Advice on infection prevention and control should be improved for people with GI symptoms, including returning travellers, to ensure hand hygiene and appropriate swimming pool avoidance.

**Data summary:** *Cryptosporidium* is a notifiable agent in the UK which diagnostic laboratories must report to local health protection teams. Submission of *Cryptosporidium*-positive stools to the CRU is voluntary, but allows characterisation of the species. We used these data, where the specimen originated from English and Welsh diagnostic laboratories, to describe the epidemiology of *Cryptosporidium* spp. between 2015 and 2021.

**Impact statement:** *Cryptosporidium* infections in industrialised countries can cause serious disease and lead to complicated and lasting sequelae, especially in the immunocompromised. Even in the general population, as well as long term gastrointestinal upset, joint pain, headache and eye pain have also been identified more frequently following cryptosporidiosis (1). There is an established association between cryptosporidiosis and colorectal cancer, although no conclusive evidence regarding causality in either direction (2–5). There has never been such a dramatic reduction in international travel in the modern era than during the COVID-19 pandemic, which is a key driver of *C. hominis* infections. Conversely, pressure on outdoor amenities has rarely been higher, which posed an increase in the likelihood of infection and cross-contamination for *C. parvum* infections. There have been few time-series analyses of cryptosporidiosis; in order to inform and strengthen surveillance programmes, we aimed to assess if there was a significant change to the epidemiology of *C. parvum* and *C. hominis* during the COVID-19 pandemic.

## Introduction

Cryptosporidiosis is a zoonotic disease caused primarily by the protozoan parasites *Cryptosporidium hominis* and *Cryptosporidium parvum*. It is most common in children aged between one and five years (6–8). People with weak immune systems, especially severe T-cell deficiencies, are usually more seriously affected (9,10). The most common symptom is mild to severe watery diarrhoea, often accompanied by abdominal cramps, nausea/vomiting, low-grade fever, weight loss and dehydration (7).

Symptoms begin three to 12 days (average five to seven days) after infection. In healthy people, symptoms usually last about one to two weeks but can persist for up to a month. The symptoms may be cyclical, where patients seem to get better for a few days, then feel worse again before the illness ends. In the immunocompromised, illness can be severe and protracted and sometimes fatal. There is an association with previous infection and developing colorectal cancer, although no causative proof (2–5). Long-term sequelae such as diarrhoea, abdominal pain, nausea, fatigue and headache are common (1) and infection can cause cognitive deficit and failure to thrive in malnourished young children in moderate-to-low income countries (11).

Diagnosis is performed by microscopy (acid-fast or fluorescent staining) or immunoassay to detect oocyst antigens or PCR to detect DNA. Genotyping by PCR is used as a reference test to differentiate species; *C. parvum* and *C. hominis* may be further sub-typed by sequencing part of the *gp60* gene (12,13) at the national Cryptosporidium Reference Unit (CRU) in Swansea, Wales. A multi-locus variable number of tandem repeats analysis (MLVA) (14) by fragment sizing has been validated and recently implemented by the CRU (15).

*C. parvum* also affects ruminants (mainly sheep and cattle in the UK) and is zoonotic while *C. hominis* is predominantly anthroponotic (transmitted directly or indirectly from person-to-person) (6–8). Household transmission is important, especially for *C. hominis* (8). Outbreaks of cryptosporidiosis have been linked to drinking or swimming in and ingesting contaminated water, contact with infected lambs and calves during visits to open or commercial farms, person to person spread in institutional settings and consumption of contaminated food items (7,16,17). The parasites are resistant to chlorine but large enough to be captured by appropriate water filtration systems (18). The majority of outbreaks in England and Wales are linked to animal contact at open/petting farms (exclusively *C. parvum*) and swimming pools (vast majority *C. hominis*) (13).

UK cryptosporidiosis cases display a seasonal trend: a late spring peak for *C. parvum* (often associated with greater countryside activities, opening of farm-based leisure activities and the lambing season) and an early autumn peak for *C. hominis* (often associated with overseas travel and summer activities such as swimming) (19,20).

COVID-19 greatly limited foreign travel for most UK residents in 2020; official figures show a 74% reduction in visits abroad for any reason (21), whereas the previous decade had seen a steady growth in foreign travel. There was a more than 500% increase in the number of people seeking holidays or leisure activities within the UK in 2020 (22) and also an increase in the number of people using outdoor spaces in the UK in 2020-21, including walking, cycling or “wild” swimming (23–25). Furthermore, people might have undertaken these activities in areas new to them and where they were unaware of locally understood health risks. People were also more likely to wash their hands and less likely to use swimming pools and restaurants because of COVID-19 restrictions (26,27). Reductions in GI illness were observed across all surveillance indicators as COVID-19 started to peak. Compared with the 5-year average (2015–2019), there was a 52% reduction in GI outbreaks reported during the first 6 months of the COVID-19 response (28). We aimed to assess if COVID-19 restrictions caused a significant change to the epidemiology of *C. parvum* and *C. hominis*.

## Methods

### Study design

To estimate the impact of COVID-19 restrictions on *C. hominis* and *C. parvum*, we conducted a retrospective observational study using interrupted time-series analysis with generalised linear modelling.

### Study population and data source

We analysed all confirmed cases of all *Cryptosporidium* species (29) extracted from the CRU database for the period between 01 January 2015 to 31 December 2021. Date of case was defined as the date the specimen was received by the CRU. Records were imported into Stata V14 and cleaned to remove quality control specimen data and duplicate reports. We retained only cases with *C. hominis* and *C. parvum* infections who were resident in England and Wales or, if the case’s address was not known, where the specimen had been sent to the CRU from a laboratory in England and Wales. We merged the CRU data using case’s resident postcode with Office for National Statistics (ONS) data for deprivation (30) and rural/urban classification (31). We used the date of the first UK-wide lockdown on 23 March 2020 (week 13) to create a pre-restrictions period and a post restrictions-implementation period (COVID=0 between week 1, 2015 and week 12, 2020 and COVID=1 between week 13, 2020 and week 52, 2021).

### Statistical analysis

We stratified results by age, sex, deprivation, rural/urban assignment and foreign travel. Foreign travel is often poorly recorded on stool specimen forms. Those with foreign travel indicated are considered reliable data by the CRU but those with “no”, “null” or missing are not considered reliable. Here, proportion with foreign travel was calculated as total indicating yes to foreign travel for that subgroup, divided by the total number of cases. Time series analysis uses regression methods to illustrate trends in the data. It incorporates information from past observations and past errors in those observations into the estimation of predicted values (32–34). We conducted an interrupted time-series analysis for *C. hominis* and *C. parvum* cases using negative binomial regression to account for over dispersion and secular trend. Models included linear splines to test for differences in incidence between the pre- and post-restriction periods, and Fourier analysis to adjust for underlying periodicity. Selection of the final model was informed by the Akaike information criterion (AIC)(35).

A separate model, following the same principles as above, was built for *C. hominis* and *C. parvum* cases up to week 13, 2020 and then extended to week 52, 2021 to forecast the number of cases expected had COVID-19 not occurred.

## Results

### Descriptive epidemiology

There were 21,304 cases between 01 January 2015 to 31 December 2021 (*C. hominis*=9,058; *C. parvum*=12,246). There were 8,991 cases of *C. hominis* pre-restrictions and 67 post restrictions-implementation compared to 9,732 cases of *C. parvum* pre-restrictions and 2,514 post restrictions-implementation. For *C. hominis*, weekly case-total variation was between zero to 180 cases pre-restrictions and zero to four cases post restrictions-implementation. For *C. parvum*, the range was zero to 162 pre-restrictions and six to 63 post restrictions-implementation. A periodicity of 52 weeks was observed for *C. hominis* and of 26 and 52 weeks for *C. parvum*.

Age data was complete but sex classification was unknown for 0.4% of *C. hominis* (n=35) and 0.2% (n=29) of *C. parvum* cases. Age distribution was similar pre- and post-restrictions for *C. hominis* (SI figure 1) and *C. parvum* (SI figure 2), with children aged 0-9 years experiencing a higher proportion of cases.

Cases of *C. hominis* were more likely to be female pre-restrictions but male post restrictions-implementation(SI figures 3). For *C. parvum*, cases were more likely to be female pre-restrictions and post restrictions-implementation (SI figure 4).

Postcodes were not recorded for approximately 10% of cases (*C. hominis* n=858; *C. parvum* n=1,139), meaning that deprivation and rural/urban ranking could not be assigned. There was a social gradient observed for *C. hominis* cases pre-restrictions, where the proportion of cases increased with deprivation. This social gradient didn’t remain post restrictions-implementation but there was a large increase in the proportion of cases from the most deprived decile (SI figure 5), where 27.2% of *C. hominis* cases were from the most deprived decile compared to 12.7% pre-restrictions. For *C. parvum* cases, the social gradient was the inverse of *C. hominis* but remained stable pre- and post restrictions-implementation, with at 7.6% and 6.7% respectively in the most deprived decile (SI figure 6).

Cases were more likely to live in an urban area for both species pre-restrictions and post restrictions-implementation. The protective nature of rural residence was more pronounced for *C. hominis* cases (SI figure 7) than for *C. parvum* cases (SI figure 8). Foreign travel status was missing for most cases (*C. hominis* missing=65%, n=5,865; *C. parvum* missing=75%, n=9,137). Where recorded, 65% of *C. hominis* cases (n=1,980 pre-restrictions [63%]; n=22 post restrictions-implementation [76%]) and 33% of *C. parvum* cases (n=970 pre-restrictions [34%]; n=41 post restrictions-implementation [14%]) had travelled abroad. For *C. hominis* cases, the most common travel destinations pre-restrictions were Spain (n=296), Turkey (n=149), Pakistan (n=135), India (n=124) and Egypt (n=98). These cases were distributed fairly evenly across all deprivation deciles with the exception of Pakistan where 67% were from cases in deciles one to three. Post restrictions-implementation, 22 *C. hominis* cases had travelled to Pakistan and half of these cases were in deprivation deciles one to three. For *C. parvum* cases, the most common travel destinations pre-restrictions were Portugal (n=69), Spain (n=65), Turkey (n=64), France (n=63), Pakistan (n=51) and India (n=35). These cases were again distributed fairly evenly across all deprivation deciles with the exception of those who had been to Pakistan, with 63% (n=32) of cases ranked in deciles one to three.

### Time series analysis

For both species, our final TSA model was improved by using Fourier transforms for 26 week and 52 week periodicity, based on the AIC test results. A negative binomial regression model provided a better fit than a Poisson model. The time-series data for *C. hominis* had a main periodicity of 52 weeks, with annual peaks clearly visible in autumn. An interaction was found between our COVID variable and both Fourier waves so these were retained in our final model. Comparing the two models (full dataset and forecasted data based on the previous five years’ data to predict cases had COVID-19 not occurred) highlighted the extreme reduction of *C. hominis* cases post restrictions-implementation (figure 1).

**Fig 1.**
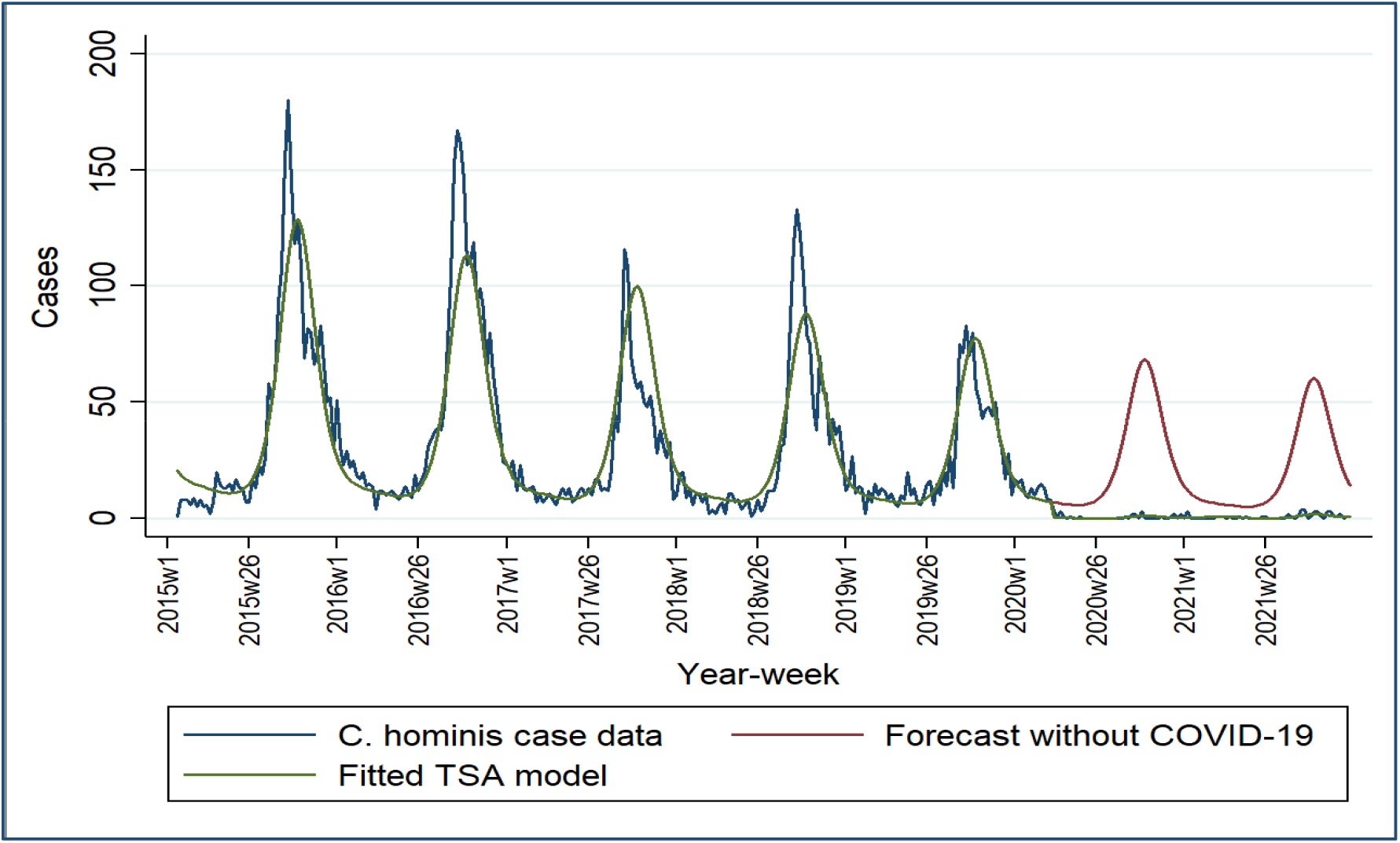
*C. hominis* model including forecast had COVID-19 not occurred, England & Wales, 2015-2021.

Post restrictions-implementation, the incidence of *C. hominis* dropped by 97.5% (95%CI: 95.4%-98.6%; p<0.001) (table 1). A decreasing incidence-trend pre-restrictions (n=8,991; IRR=0.9976; 95%CI: 0.9969-0.9982; p<0.001) of 0.24% reduction in cases per week was not observed post restrictions-implementation (n=67; IRR=1.0081; 95%CI: 0.9978-1.0186; p=0.128). Whilst a periodicity change was observed post restrictions-implementation for *C. hominis* (based on the COVID#c.sin52 interaction term; table 1), the forecast model did not show a change in peak incidence by week number (week 41).

**Table 1.** Time-series analysis model results for *C. hominis*.

| *C. hominis* TSA model | IRR | p-value | 95% CI |
| --- | --- | --- | --- |
| Pre COVID period | 0.997579 | <0.001 | (0.9969615, 0.9981975) |
| Post COVID period | 1.0081 | 0.128 | (0.9976892, 1.01862) |
| Post vs. pre period | 0.024829 | <0.001 | (0.0135401, 0.0455293) |
| sin52* | 0.308177 | <0.001 | (0.2878808, 0.329904) |
| COVID#c.sin52<sup>L</sup> | 2.616494 | <0.001 | (1.822834, 3.755715) |
| cos52* | 1.291586 | <0.001 | (1.204945, 1.384458) |
| COVID#c.cos52<sup>L</sup> | 1.279372 | 0.321 | (0.7864356, 2.081279) |
| sin26* | 0.994242 | 0.867 | (0.9295001, 1.063493) |
| COVID#c.sin26<sup>⊥</sup> | 1.026261 | 0.899 | (0.6880788, 1.530657) |
| cos26* | 0.732617 | <0.001 | (0.6842376, 0.784418) |
| COVID#c.cos26<sup>⊥</sup> | 0.674863 | 0.057 | (0.4503512, 1.0113) |

\*sin26&cos26 and sin52&cos52 are Fourier transforms. These are part of the same waveform. Interaction terms measuring the effect of COVID-19 on the periodicity of cases.

The time-series data for *C. parvum* had a periodicity of 26 and 52 weeks, with biannual peaks visible in spring and in autumn. An interaction was found between our COVID variable and both Fourier waves so these were retained in our final model. In the forecast model of the previous five years’ time-series data beyond week 13, 2020 for *C. parvum*, we observed a larger number of cases than predicted had COVID-19 not occurred, but not in the same order of magnitude as predicted for *C. hominis* (figure 2). Incidence of *C. parvum* had recovered to pre-restrictions levels by the end of 2021.

**Fig 2.**
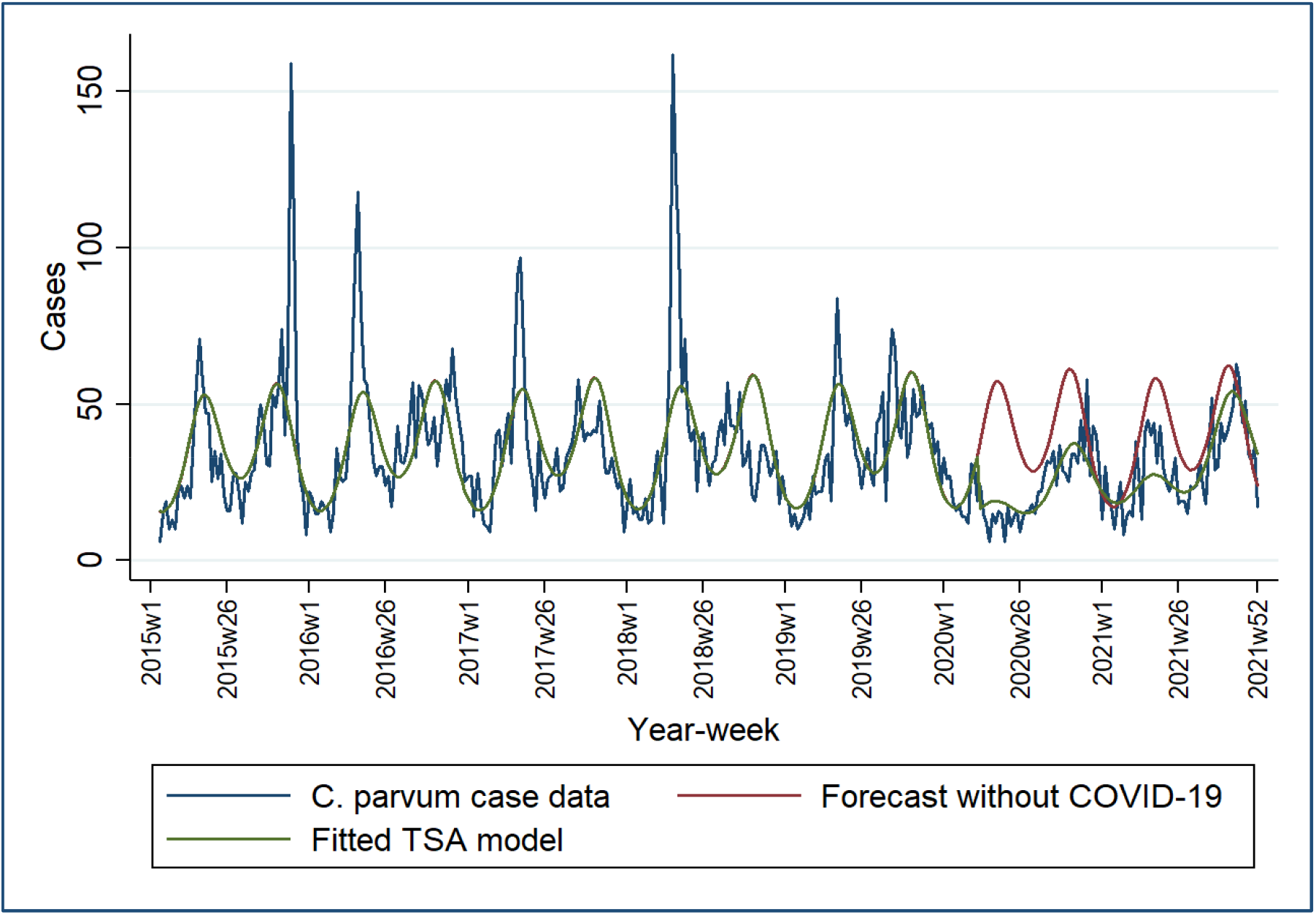
*C. parvum* model including forecast had COVID-19 not occurred, England and Wales, 2015-2021.

Post restrictions-implementationincidence of C. parvum dropped by 49.0% (95%CI: 38.4%-58.3%; p<0.001) (table 2). There was no pre-restrictions incidence-trend (n=9,732; IRR=1.0003; 95%CI: 0.9997-1.0009; p=0.322) but a slight increasing incidence-trend existed post restrictions-implementation of 0.71% increase in cases per week (n=2,514; IRR=1.0071; 95%CI: 1.0038-1.0104; p=0.001). A periodicity change was observed for C. parvum post restrictions-implementation, peaking one week earlier in spring (week 18 as opposed to week 19) and two weeks later in autumn (week 44 as opposed to week 42) (figure 3).

**Fig 3.**
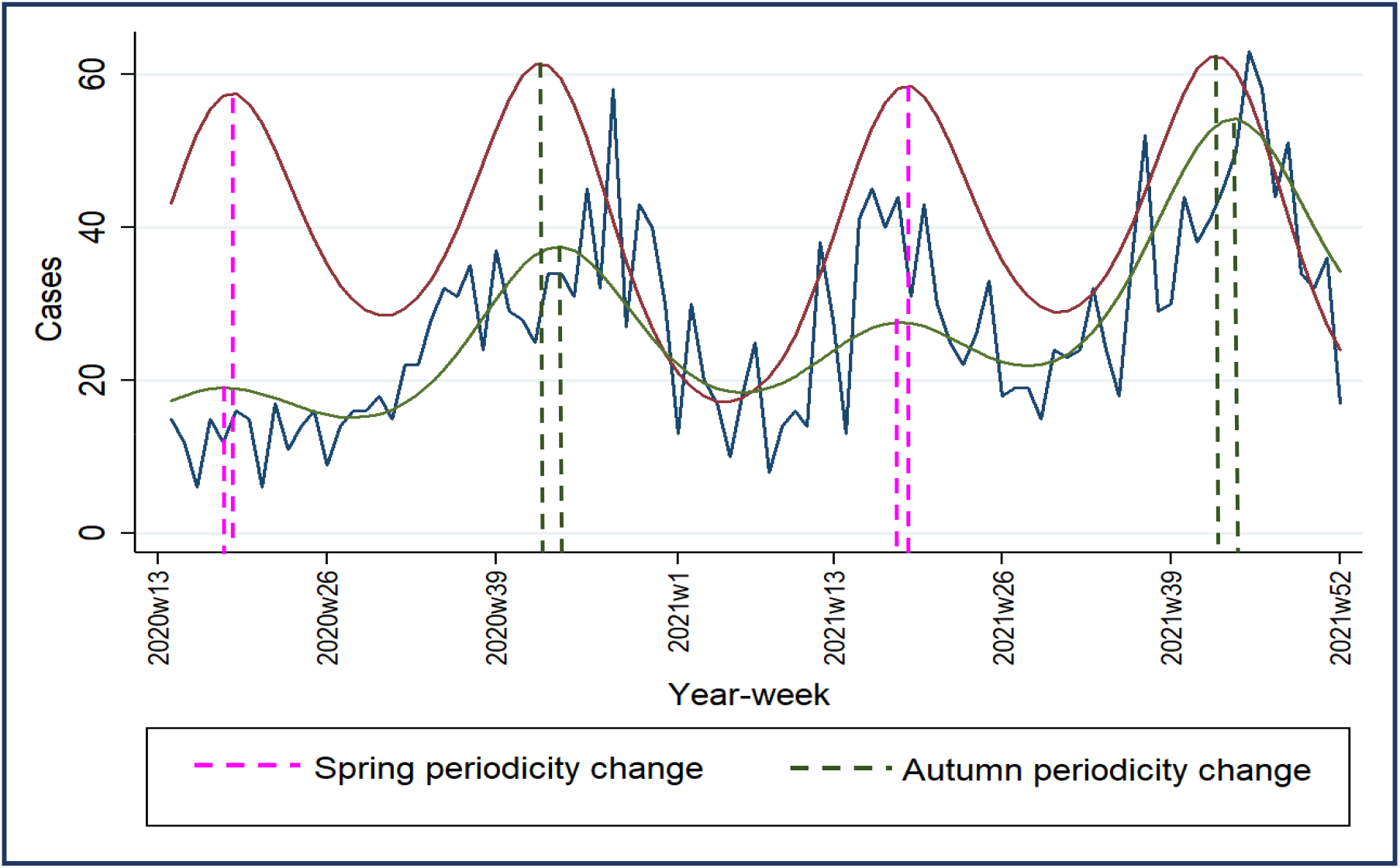
*C. parvum* model including forecast had COVID-19 not occurred and spring and autumn periodicity changes, England and Wales, 2020 week 13 to 2021 week 52.

**Table 2.** Time-series analysis model results for *C. parvum*.

| *C. parvum* TSA model | IRR | p-value | 95% CI |
| --- | --- | --- | --- |
| Pre COVID period | 1.000301 | 0.322 | (0.9997063, 1.000895) |
| Post COVID period | 1.007118 | <0.001 | (1.003844, 1.010402) |
| Post vs. pre period | 0.5067641 | <0.001 | (0.4169558, 0.6159162) |
| sin52* | 0.8763961 | <0.001 | (0.8227561, 0.9335333) |
| COVID#c.sin52<sup>L</sup> | 0.9076778 | 0.153 | (0.7946875, 1.036733) |
| cos52* | 0.801548 | <0.001 | (0.7508416, 0.8556787) |
| COVID#c.cos52<sup>L</sup> | 1.367283 | <0.001 | (1.191219, 1.569369) |
| sin26* | 0.6928407 | <0.001 | (0.6505466, 0.7378845) |
| COVID#c.sin26<sup>L</sup> | 1.186179 | 0.012 | (1.037695, 1.355909) |
| cos26* | 0.7253185 | <0.001 | (0.679221, 0.7745444) |
| COVID#c.cos26<sup>L</sup> | 1.153636 | 0.033 | (1.011625, 1.315583) |

\*sin26&cos26 and sin52&cos52 are Fourier transforms. These are part of the same waveform. Interaction terms measuring the effect of COVID-19 on the periodicity of cases

## Discussion

COVID-19 restrictions had a significant effect on the number of cases of gastrointestinal illness (GI) in England and Wales including *Cryptosporidium*(28), and in this study we have identified that they impacted both *C. hominis* and *C. parvum*. Whilst *C. parvum* cases were reduced by around half, *C. hominis* cases were almost entirely arrested. Whilst our model for *C. hominis* detected a change in periodicity (table 1: COVID#c.sin52 IRR=2.616494, 95%CI 1.822834-3.755715; p<0.001), inspection of the pre- and post restrictions-implementation modelled-data showed the autumn peak occurred at the same point in 2020 and 2021 as in the preceding five years (week 41). With just 67 cases in the entire 62-week post restrictions-implementation period, the impact of small numbers effected the precision of our model. Where we would expect tens of cases in each week, there were just a handful and often none at all post restrictions-implementation. Had these samples been submitted to the CRU a few days either side of the date received, the results could have swung significantly in either direction.

It is not possible to quantify exactly the extent to which COVID-19 restrictions reduced social mixing or any commensurate effect on person-to-person transmission, which is a limitation. *C. hominis cases* had been declining in the five years leading to COVID, despite foreign travel increasing. It is possible that other interventions were influencing these data, such as improved compliance and awareness in swimming pool filtration and avoidance with GI symptoms. We do know that public venues were closed for much of our post restrictions-implementation period, including known sources of *Cryptosporidium* infection such as open/petting farms, restaurants and swimming pools. Similarly, we cannot quantify how much use of the countryside increased post restrictions-implementation regardless of UK-based holiday figures, although anecdotally there were many news stories covering this phenomenon, particularly involving parties and swimming in rivers. Despite the large reduction in international travel post restrictions-implementation, there remains a strong association between infection and travel abroad, especially for *C. hominis*, which confirms other studies’ findings that it is a principal driver of new infections in England and Wales (19,36). More should be done to prevent returning travellers with GI symptoms infecting others through improved IPC advice about hand hygiene and exclusion / voluntary avoidance from swimming pools until two weeks after symptoms have resolved. This itself relies on people seeking healthcare with GI symptoms.

Another limitation in our analysis is the effect that reduced healthcare provision and health-seeking behaviours had on specimen submission post restrictions-implementation, particularly at the start of the COVID-19 pandemic. There is international evidence showing that people sought help less often, and those who were only mildly ill did not seek help at all (37). There was genuine fear about being infected with SARS-CoV-2 in health care settings early on when lack of an effective vaccination or definitive therapy were at the forefront of many people’s minds (38,39). This might have attenuated people’s opinion of diarrhoea and vomiting symptoms, or decisions to seek medical care. Likewise, there might have been disruptions in access to, and application of, the stool specimen and diagnostic process, resulting in fewer cases being detected.

The fact that people living in the most deprived areas experienced the highest proportion of *C. hominis* cases both pre-restrictions and post restrictions-implementation highlights potential opportunities for public health interventions. Of those in the most deprived decile where travel information was available post restrictions-implementation, 83% had surnames suggestive of a non-white ethnic background (n=5). This, coupled with the fact that in 36% of all cases where there had been foreign travel involved Pakistan, provides a compelling opportunity to target interventions to outgoing and returning travellers. However, the limited data available for travel history and ethnicity means we cannot draw absolute conclusions, despite the length of our study period. Ethnicity is often poorly reported for many diseases and this reduces the ability to understand the true impact of cryptosporidiosis in specific communities. Promotion of accurate ascertainment of ethnicity and travel history should be sought at the point of specimen collection to improve future surveillance. This would require a change to the sample submission form sent to the laboratory and training for clinicians taking history. The fact that cases of both species were more likely to live in an urban residence should be interpreted with caution. Our data, being laboratory-based, did not include details of recent trips to the countryside, occupation or contact with animals and its relevance is diminished further where foreign travel is indicated.

Our results also demonstrate the need to improve rates of specimen submission to the CRU where there is currently under-representation, especially from areas that serve large clusters of ethnic minority communities. A submission bias exists in that of the approximately 4,500 human *Cryptosporidium* cases a year in England and Wales (40), only around half are submitted to the CRU for genotyping on average. This proportion is much higher in Wales and the north west of England, while some parts of England (notably London, the South East, South West and parts of Yorkshire) are under-represented. Diagnostic laboratories have been reminded by the CRU in May 2022 to send *Cryptosporidium* positive stools for genotyping, and that the service is free to users.

This was a natural experiment which reinforced existing knowledge about transmission pathways for *C. hominis* and *C. parvum*. It also provides insight into where to focus public health efforts to reduce the risk of infection and increase the consistency of specimen submission to the CRU. National surveillance was historically of the genus *Cryptosporidium*, without species identification. Although the CRU has been genotyping from *Cryptosporidium*-positive stools submitted by laboratories throughout England and Wales since 2000, capture of these data by the UK Health Security Agency’s (UKHSA) “second-generation surveillance system” (SGSS) database commenced in 2015. These show the location of infection, case-demographics and species typing. This systematic surveillance of species typing allows outbreaks to be more clearly delineated and can track the spread of *C. parvum* as well as *C. hominis* in humans. This, and additional subtyping by gp60 sequencing, can also improve outbreak management by indicating possible exposures and strengthen epidemiological links (13). Multi-locus genotyping, or next generation sequencing (NGS), can add further detail about the genetic relationship between cases and the plausibility of infection sources (14); an MLVA scheme has been implemented for C. parvum, following validation (15) and a pilot in 2021(41) and a process for the consideration of NGS for *Cryptosporidium* surveillance and outbreaks has been commenced.

Most gastrointestinal infections reduced dramatically during the COVID-19 pandemic (37) as childcare and educational settings were closed and eating establishments’ opening hours and social / leisure activities were curtailed. Our hypothesis was that the COVID-19 restrictions might have altered the epidemiology of cryptosporidiosis by infecting species and by time, place and person. This hypothesis was upheld insofar as we saw a massive reduction in *C. hominis* cases (likely due to reduced foreign travel, reduced person-to-person contact, swimming pool closure and increased hand hygiene) and observed *C. parvum* cases decline sharply then gradually return to pre-restrictions levels by the end of 2021 (mirroring relaxation in restrictions and more participation in outdoor activities). We did not analyse case exposure data because in many areas this was not collected during the post restrictions-implementation period; at the time of writing is still not routinely collected consistently throughout England and Wales. Public health practice has varied historically in relation to *Cryptosporidium* cases data collection (42). For example, official public health guidance for the management of cryptosporidiosis cases still refers to the “Standard Gastrointestinal Disease Questionnaire” (SI figure 9). Whilst this template contains the necessary fields to collect the required case information to document cases of cryptosporidiosis (43), it is almost certainly no longer widely used in paper format. This surveillance form template has not been updated since 2004 and the ethnicity categories no longer match the current ONS designations. It would be a worthwhile exercise to investigate how case reports are made, the questions used and how these are recorded.

## Recommendations

1. Future exceedance reporting for *C. hominis* should exclude the post restrictions-implementation period but retain it for *C. parvum* (except first six weeks post restrictions-implementation)
2. Public health advice should be given to people travelling abroad about generic gastrointestinal pathogen infection prevention and control (particularly handwashing, water consumption and food hygiene) to reduce the number of cases and person-to-person spread. Returning travellers with GI symptoms should have IPC advice about hand hygiene and swimming pool avoidance to reduce onward transmission
3. Regions with low sample-referral rates to the CRU should be encouraged to increase their submission rate in order to better quantify infections by region and to help ascertain the impacts on their population.
4. Investigate how case details are recorded by laboratories in each region and encourage better recording of travel and ethnicity data at the point of sample collection. This will help better understand the full impact of imported cases of *C. hominis* and decide when, where and how best to target public health interventions. This would serve as a quality improvement project for other GI diseases.
5. Specific research should be conducted in deprived and ethnic minority communities in relation to activities and habits at home and whilst abroad. Findings should be used to tailor public health messaging about how to reduce risk of infection whilst away and on return home.
6. Repeat a time-series analysis for CRU data (ideally every few years) to assess the impact of the end in COVID-19 restrictions, recovery of public health systems, and any changes to the epidemiology of cryptosporidiosis by species, time, place and person.

## Data Availability

All data produced in the present study are available upon reasonable request to the authors

## Nomenclature

AIC: Akaike information criterion
*C. hominis*: *Cryptosporidium hominis*
*C. parvum*: *Cryptosporidium parvum*
CI: confidence interval
GI: gastrointestinal illness
IPC: infection prevention and control
IRR: incidence rate ratio
PCR: polymerase chain reaction
CRU: Cryptosporidium Reference Unit
MLVA: multi-locus variable number of tandem repeats analysis
UK: United Kingdom
TSA: time series analysis

## Author contributions

James P. Adamson undertook the data cleaning, ran the time-series analysis, undertook statistical tests, produced the tables and figure, and drafted and edited the manuscript. Rachel Chalmers helped set the research question, provided background on *Cryptosporidium* spp., helped resolve data queries and helped edit the manuscript.

Kristin Elwin and Guy Robinson developed, validated and ran the laboratory processes that produced the typing results at the CRU and provided background on *Cryptosporidium* spp.

Daniel Thomas critically appraised the manuscript, helped to redraft and format it, and provided project supervision and quality assurance.

Alicia Barrasa supervised the time-series analysis, advising with coding and providing script quality assurance support. She also critically appraised the manuscript and provided project supervision.

All authors read the manuscript and suggested various revisions.

## Acknowledgements

Heather Ayres, Lead Biomedical Scientist, Cryptosporidium Reference Unit, Public Health Wales, Swansea, UK for specimen management and PCR testing.

Jonathan Goss, Biomedical Support Worker, Cryptosporidium Reference Unit, Public Health Wales, Swansea, UK for specimen reception and DNA extraction.

## Ethics

Ethical oversight of the project was provided by the Public Health Wales Research and Development Division. As this work was carried out using routinely collected surveillance data, PHW Research and Development Division advised that NHS research ethics approval was not required. Data were held and processed under PHW’s information governance arrangements, in compliance with the Data Protection Act, Caldicott Principles and PHW guidance on the release of small numbers. No data identifying protected characteristics of an individual were released outside of the project team.

## Data Availability Statement

The data used in this analysis contained personal identifiable information. Anonymised information, including that contained in the supplementary information, is reported by Public Health Wales and the UKHSA and published on the UK Government website. Datasets and coding scripts required to reproduce these results are available from the corresponding author on reasonable request.

## Supplementary information

**SI Figure 1.**
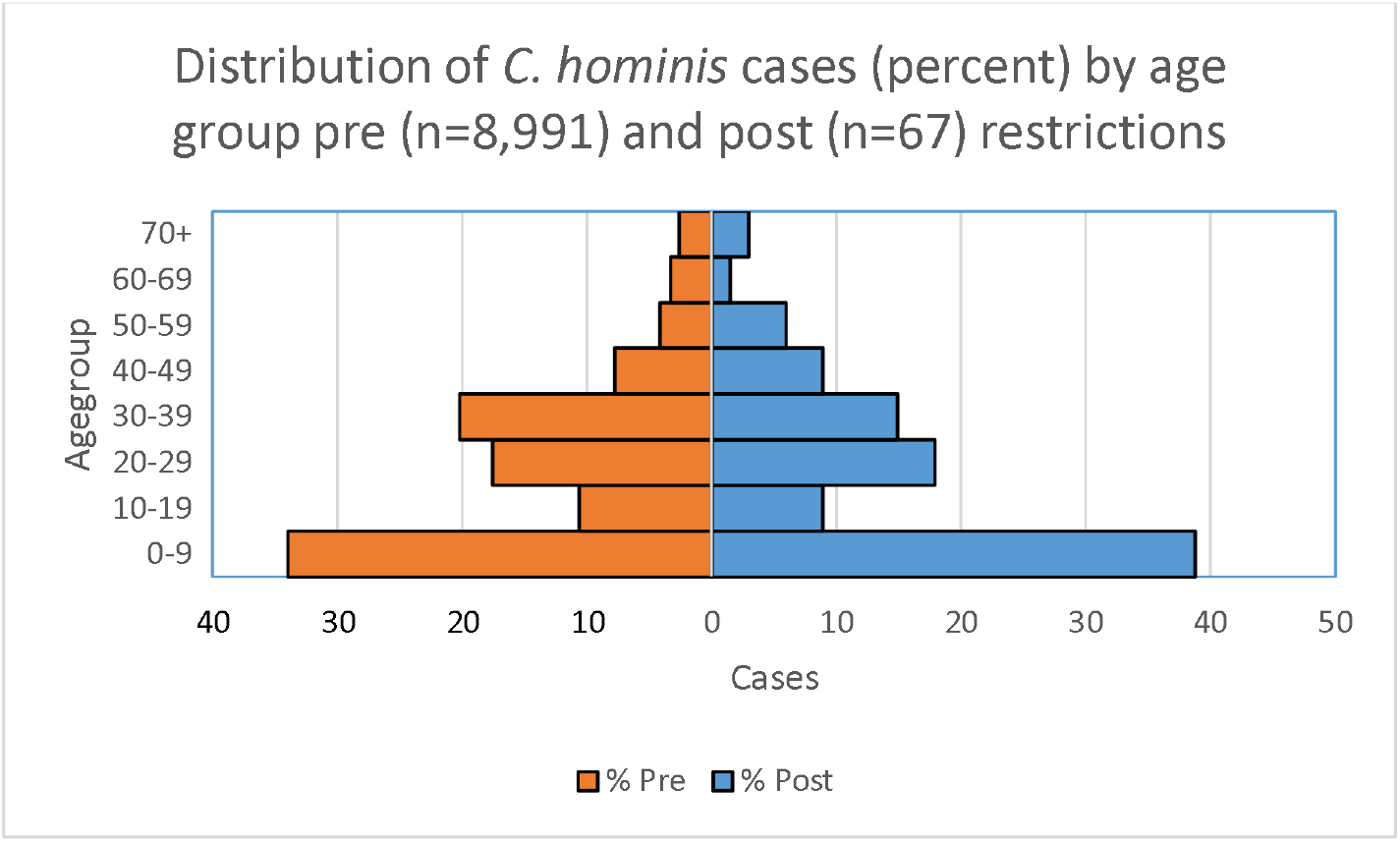
Distribution of *C. hominis* cases (percent) by age group pre (n=8,991) and post (n=67) restrictions-implementation.

**SI Figure 2.**
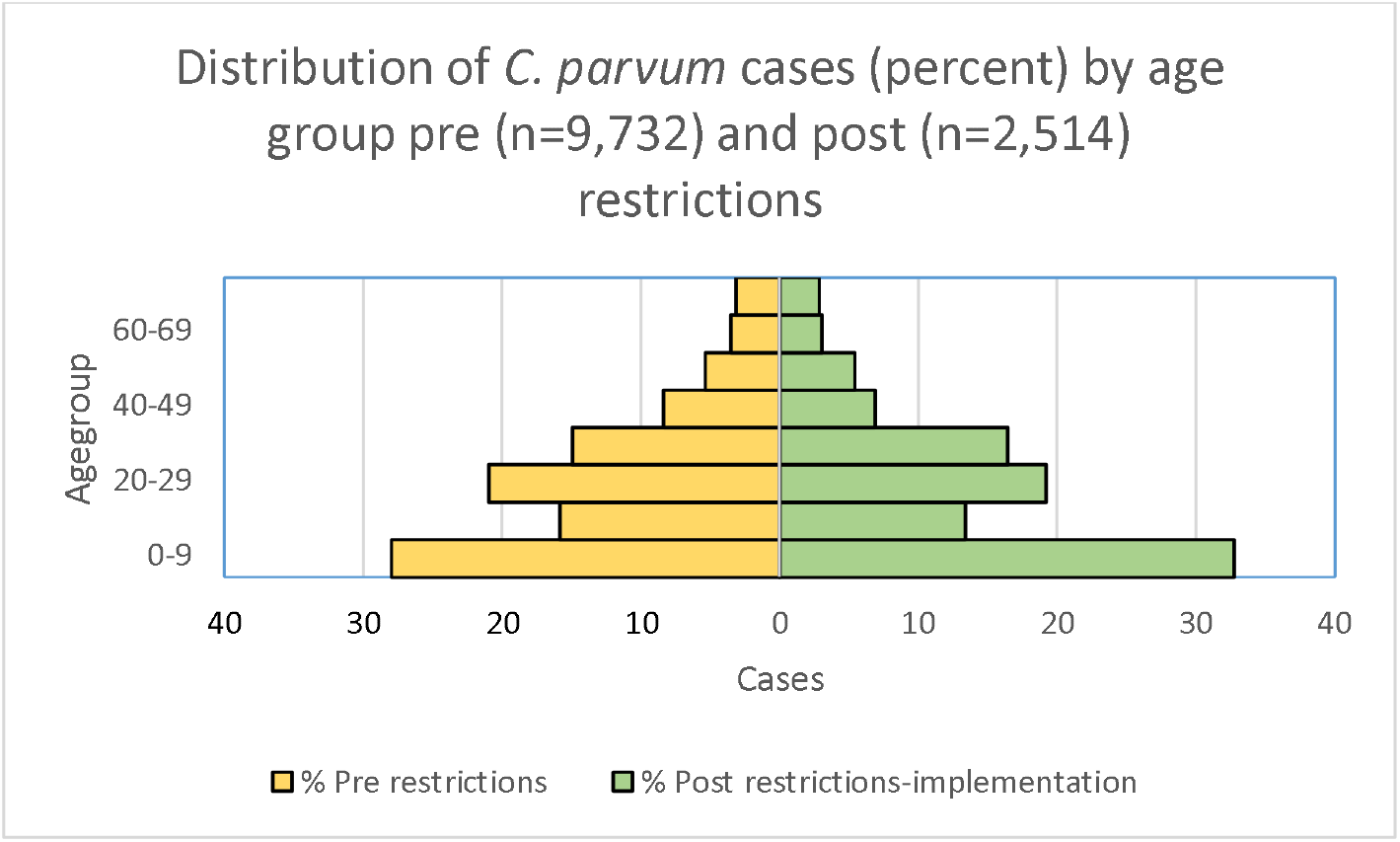
Distribution of *C. parvum* cases (percent) by age group pre (n=9,732) and post (n=2,514) restrictions-implementation.

**SI Figure 3.**
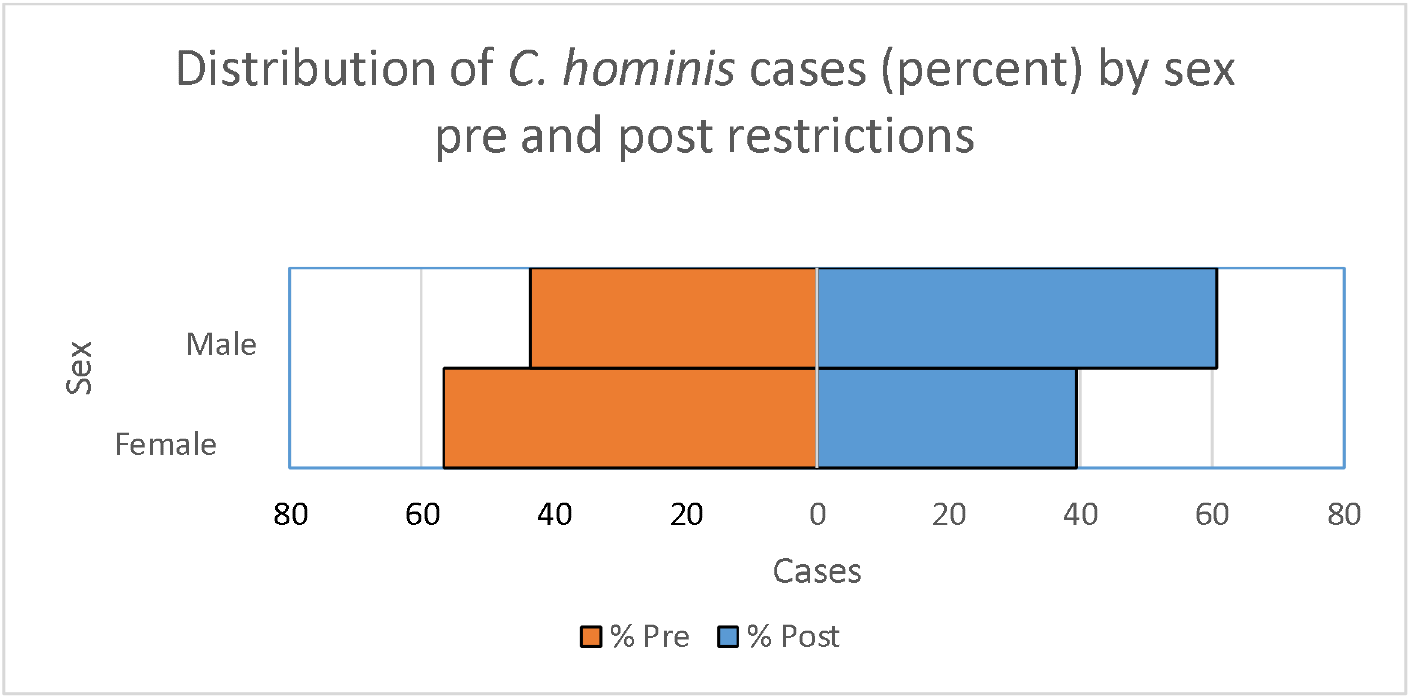
Distribution of *C. hominis* cases (percent) by sex pre (n=8,957) and post (n=66) restrictions-implementation.

**SI Figure 4.**
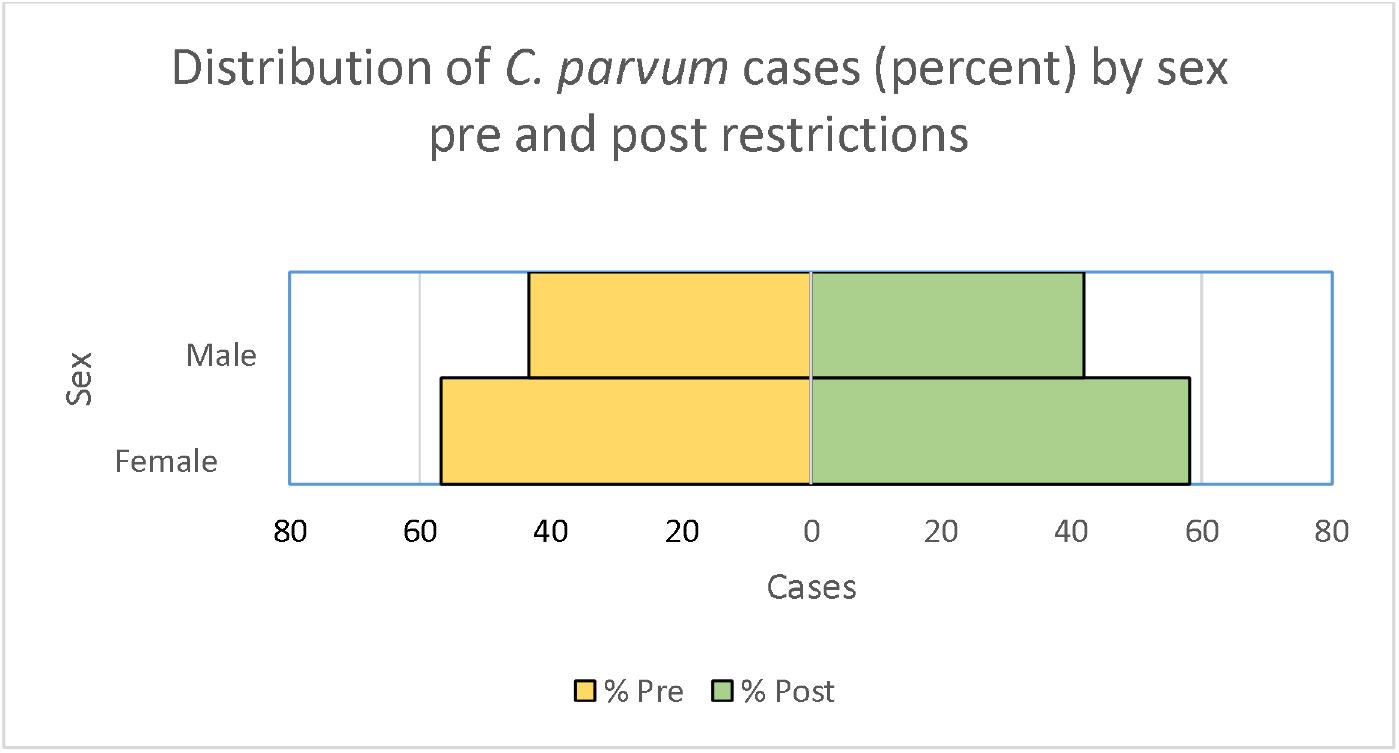
Distribution of *C. parvum* cases (percent) by sex pre (n=9,710) and post (n=2,507) restrictions-implementation.

**SI Figure 5.**
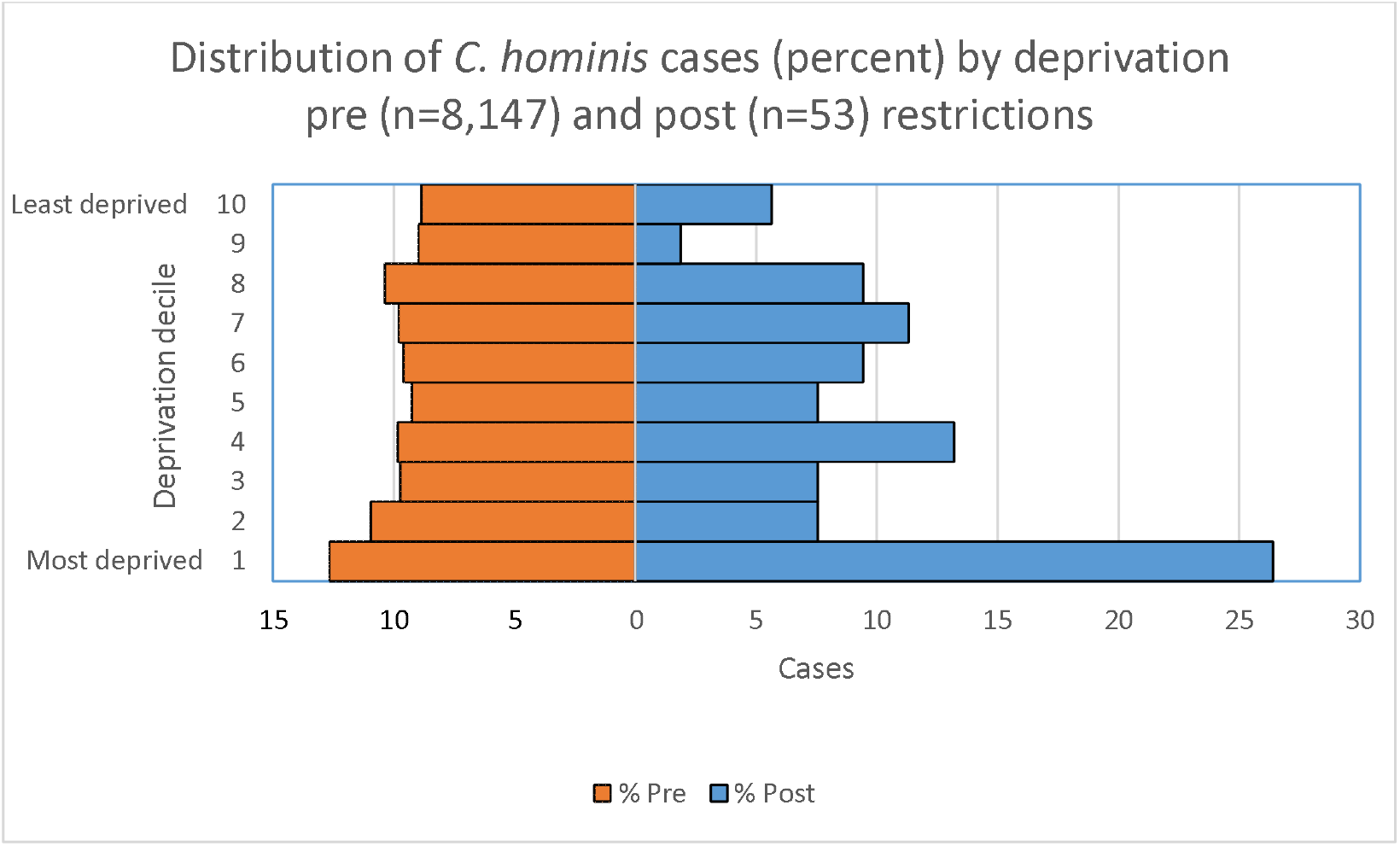
Distribution of *C. hominis* cases (percent) by deprivation pre (n=8,147) and post (n=53) restrictions-implementation.

**SI Figure 6.**
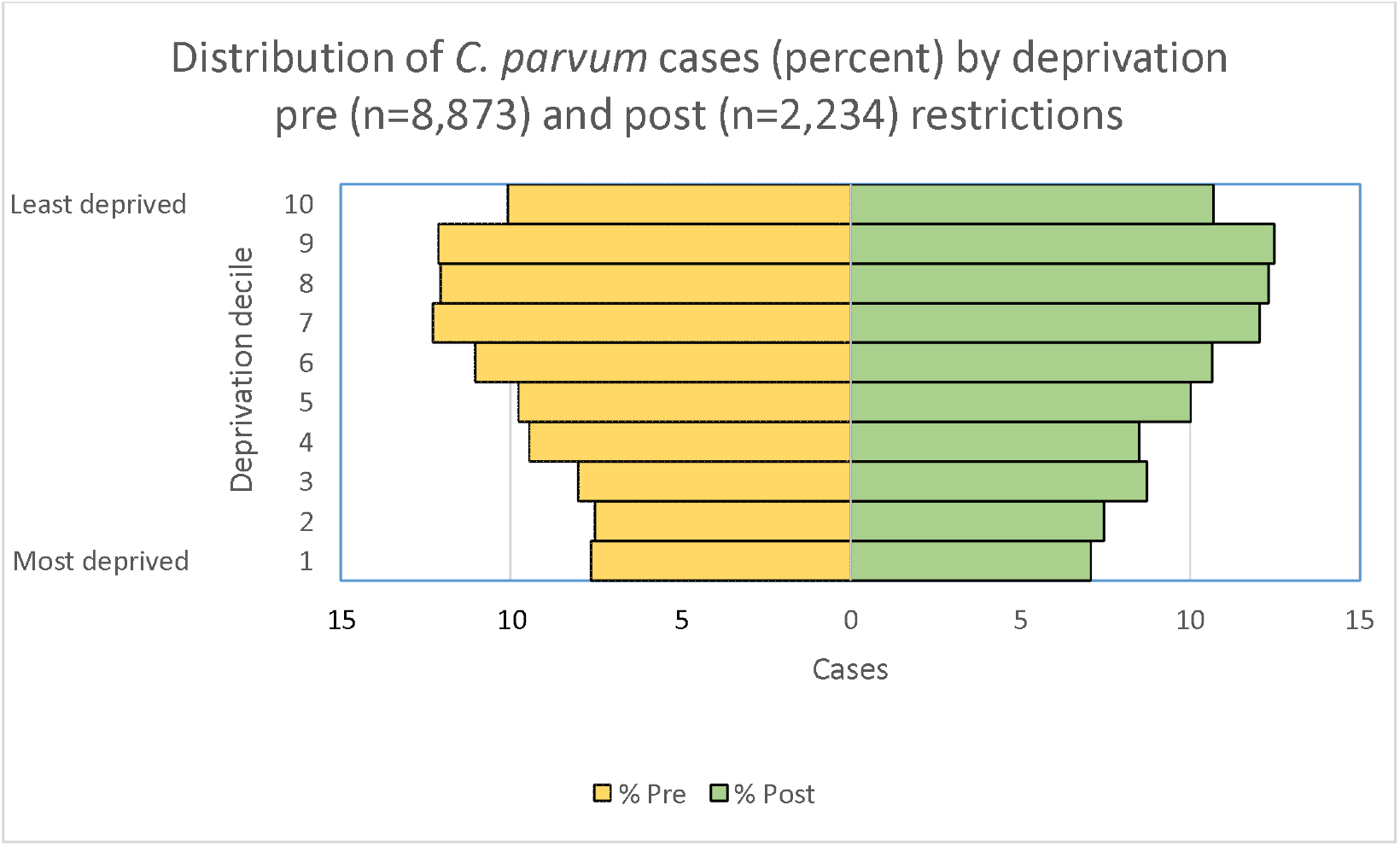
Distribution of *C. parvum* cases (percent) by deprivation pre (n=8,873) and post (n=2,234) restrictions-implementation.

**SI Figure 7.**
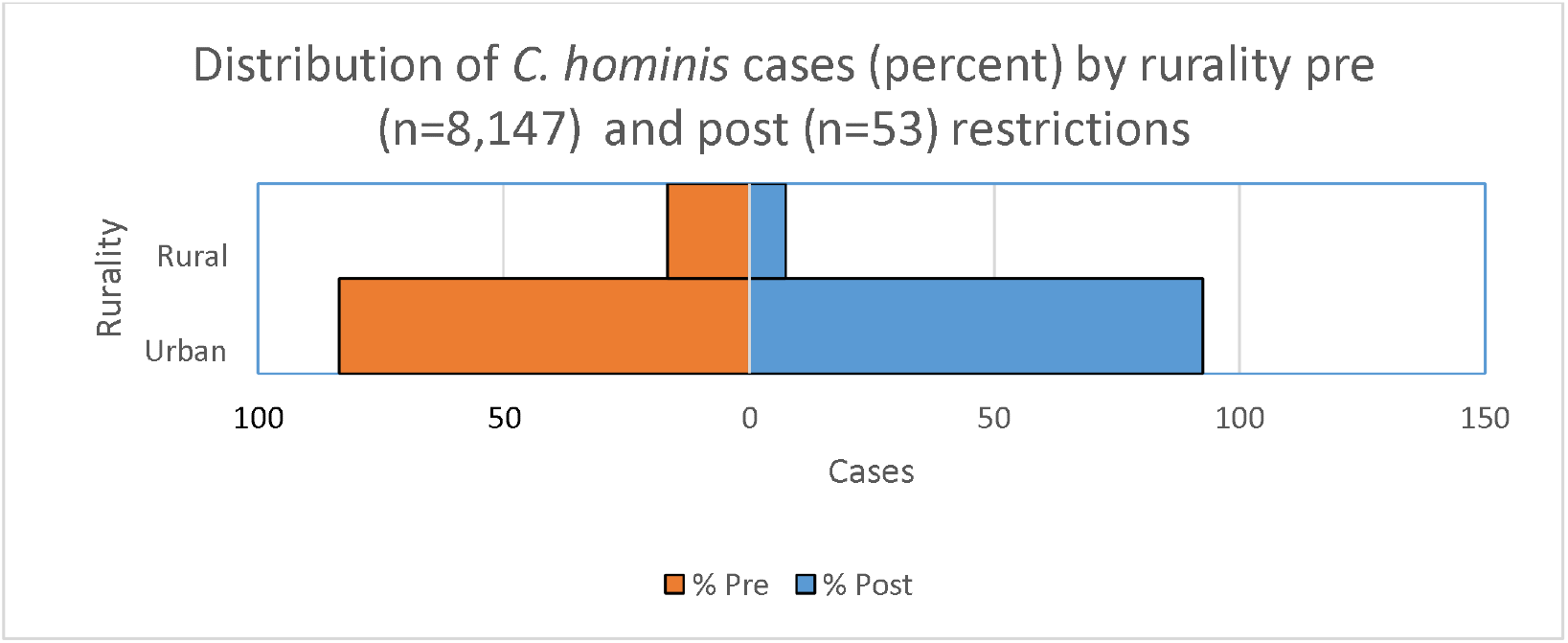
Distribution of *C. hominis* cases (percent) by residence rurality pre (n=8,147) and post (n=53) restrictions-implementation.

**SI Figure 8.**
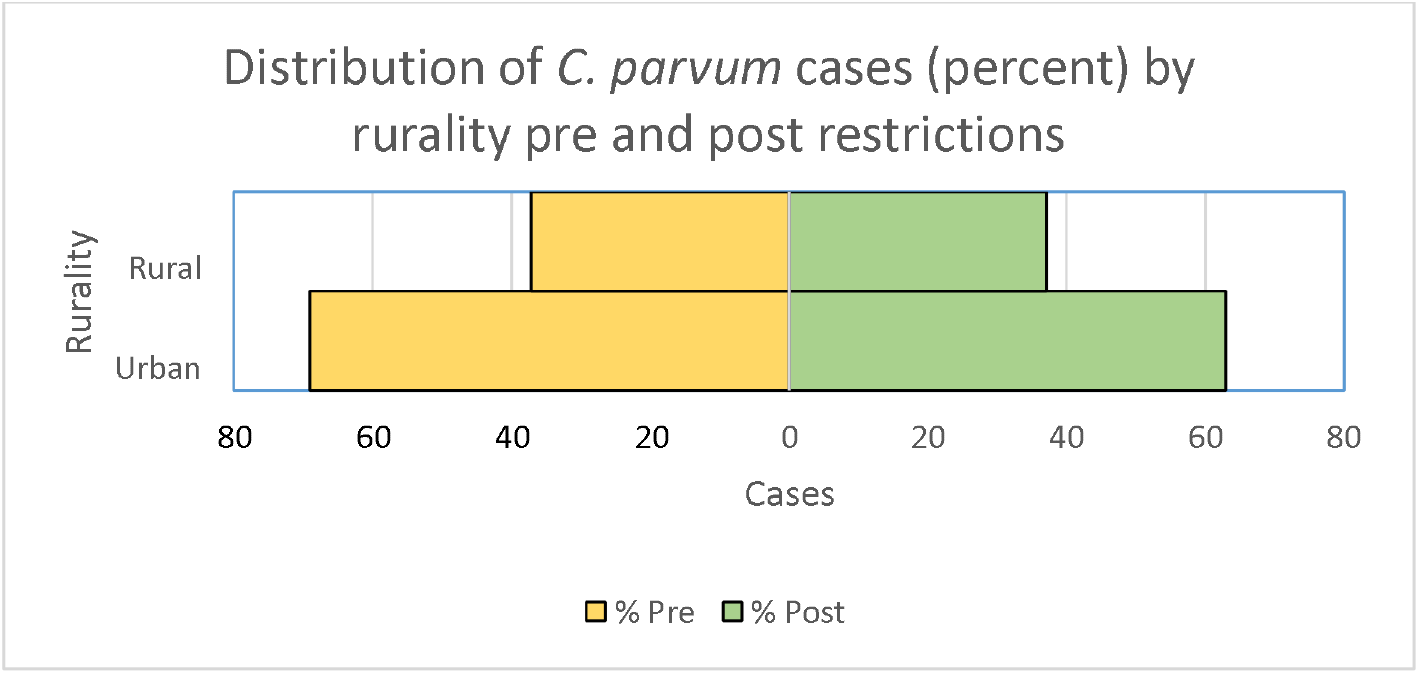
Distribution of *C. parvum* cases (percent) by residence rurality pre (n=8,873) and post (n=2,273) restrictions-implementation.

**SI Figure 9.**
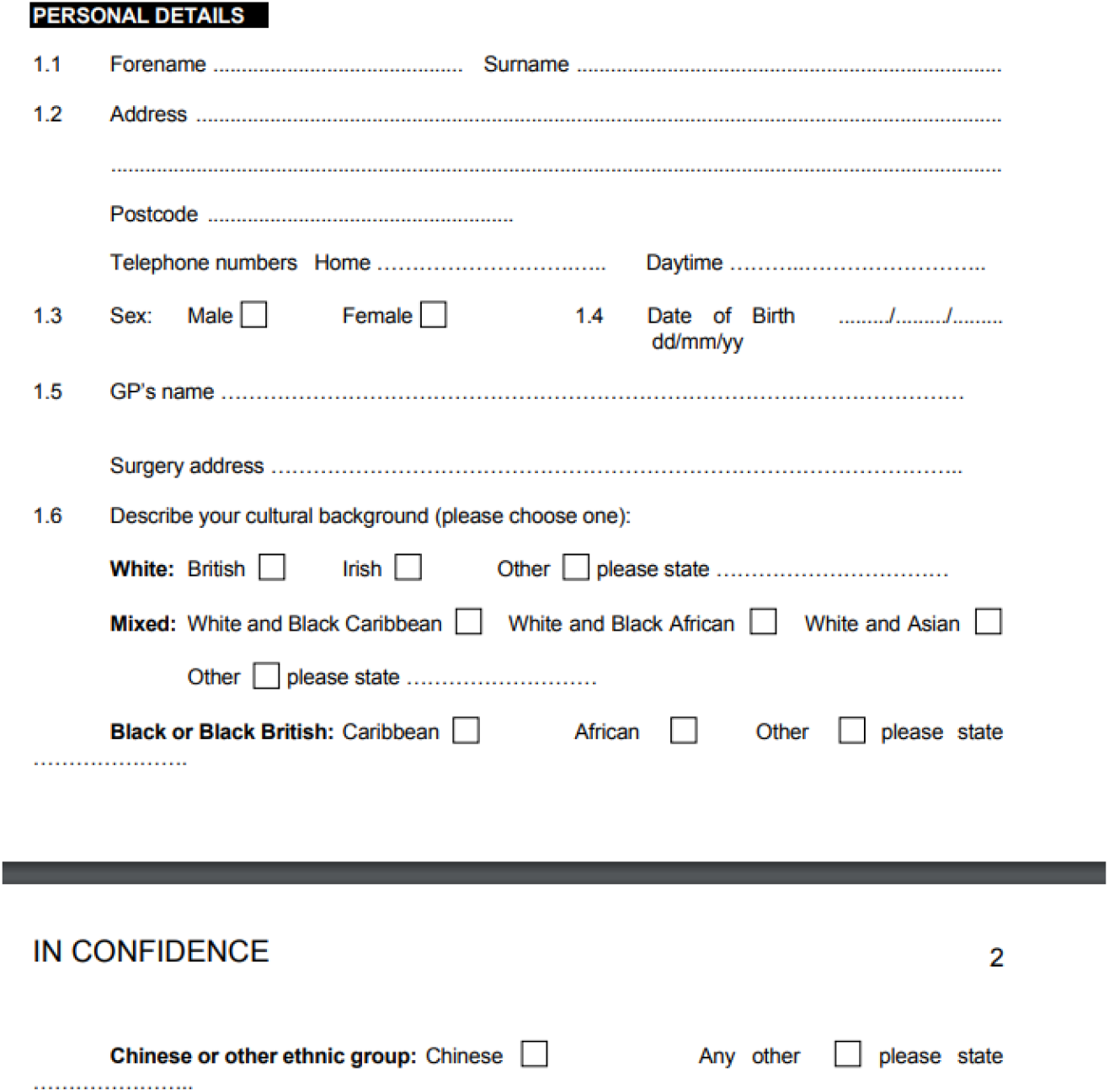
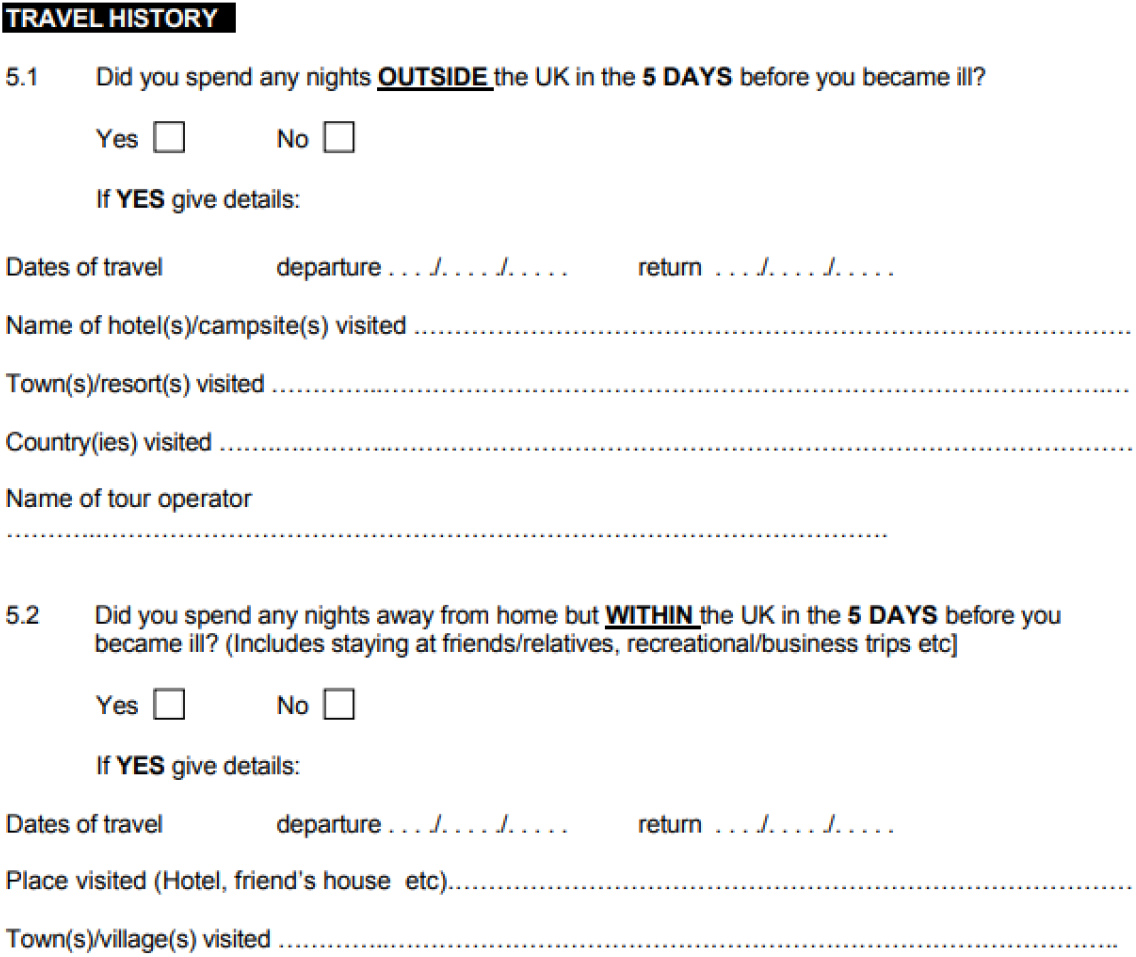
UKHSA Standard Gastrointestinal Disease Questionnaire (2004) extract containing categories for ethnicity and travel history data collection.

## References

1. Carter BL, Chalmers RM, Davies AP. Health sequelae of human cryptosporidiosis in industrialised countries: a systematic review. Parasit Vectors [Internet]. 2020;13(1):443. Available from: https://doi.org/10.1186/s13071-020-04308-7

2. Kalantari N, Gorgani-Firouzjaee T, Ghaffari S, Bayani M, Ghaffari T, Chehrazi M. Association between Cryptosporidium infection and cancer: A systematic review and meta-analysis. Parasitol Int [Internet]. 2020;74:101979. Available from: https://www.sciencedirect.com/science/article/pii/S1383576919303307

3. Sawant M, Baydoun M, Creusy C, Chabé M, Viscogliosi E, Certad G, et al. Cryptosporidium and Colon Cancer: Cause or Consequence? Microorganisms [Internet]. 2020 Oct 27;8(11):1665. Available from: https://pubmed.ncbi.nlm.nih.gov/33121099

4. Sulżyc-Bielicka V, Kołodziejczyk L, Jaczewska S, Bielicki D, Safranow K, Bielicki P, et al. Colorectal cancer and Cryptosporidium spp. infection. PLoS One [Internet]. 2018 Apr 19;13(4):e0195834–e0195834. Available from: https://pubmed.ncbi.nlm.nih.gov/29672572

5. Zhang N, Yu X, Zhang H, Cui L, Li X, Zhang X, et al. Prevalence and Genotyping of Cryptosporidium parvum in Gastrointestinal Cancer Patients. J Cancer [Internet]. 2020;11(11):3334–9. Available from: https://www.jcancer.org/v11p3334.htm

6. Perry S, Sanchez M de la L, Hurst P, Parsonnet J. Household Transmission of Gastroenteritis. Emerg Infect Dis J [Internet]. 2005;11(7):1093. Available from: https://www.nc.cdc.gov/eid/article/11/7/04-0889_article

7. Chalmers RM, Smith R, Elwin K, Clifton-Hadley FA, Giles M. Epidemiology of anthroponotic and zoonotic human cryptosporidiosis in England and Wales, 2004–2006. Epidemiol Infect [Internet]. 2010/07/12. 2011;139(5):700–12. Available from: https://www.cambridge.org/core/article/epidemiology-of-anthroponotic-and-zoonotic-human-cryptosporidiosis-in-england-and-wales-20042006/997A2C96463C3CB3F147DF6D02D026A7

8. McKerr C, Chalmers RM, Elwin K, Ayres H, Vivancos R, O’Brien SJ, et al. Cross-sectional household transmission study of Cryptosporidium shows that C. hominis infections are a key risk factor for spread. BMC Infect Dis [Internet]. 2022;22(1):114. Available from: https://doi.org/10.1186/s12879-022-07086-y

9. Abdou AG, Harba NM, Afifi AF, Elnaidany NF. Assessment of Cryptosporidium parvum infection in immunocompetent and immunocompromised mice and its role in triggering intestinal dysplasia. Int J Infect Dis [Internet]. 2013;17(8):e593–600. Available from: https://www.sciencedirect.com/science/article/pii/S1201971212013161

10. Hunter PR, Nichols G. Epidemiology and clinical features of Cryptosporidium infection in immunocompromised patients. Clin Microbiol Rev. 2002 Jan;15(1):145–54.

11. Khalil IA, Troeger C, Rao PC, Blacker BF, Brown A, Brewer TG, et al. Morbidity, mortality, and long-term consequences associated with diarrhoea from Cryptosporidium infection in children younger than 5 years: a meta-analyses study. Lancet Glob Heal. 2018 Jul;6(7):e758–68.

12. Rojas-Lopez L, Elwin K, Chalmers RM, Enemark HL, Beser J, Troell K. Development of a gp60-subtyping method for Cryptosporidium felis. Parasit Vectors [Internet]. 2020;13(1):39. Available from: https://doi.org/10.1186/s13071-020-3906-9

13. Chalmers RM, Robinson G, Elwin K, Elson R. Analysis of the Cryptosporidium spp. and gp60 subtypes linked to human outbreaks of cryptosporidiosis in England and Wales, 2009 to 2017. Parasit Vectors [Internet]. 2019;12(1):95. Available from: https://doi.org/10.1186/s13071-019-3354-6

14. Nadon CA, Trees E, Ng LK, Møller Nielsen E, Reimer A, Maxwell N, et al. Development and application of MLVA methods as a tool for inter-laboratory surveillance. Euro Surveill Bull Eur sur les Mal Transm = Eur Commun Dis Bull. 2013 Aug;18(35):20565.

15. Robinson G, Pérez-Cordón G, Hamilton C, Katzer F, Connelly L, Alexander CL, et al. Validation of a multilocus genotyping scheme for subtyping Cryptosporidium parvum for epidemiological purposes. Food Waterborne Parasitol [Internet]. 2022;27:e00151. Available from: https://www.sciencedirect.com/science/article/pii/S2405676622000087

16. Lake IR, Harrison FCD, Chalmers RM, Bentham G, Nichols G, Hunter PR, et al. Case-control study of environmental and social factors influencing cryptosporidiosis. Eur J Epidemiol [Internet]. 2007;22(11):805. Available from: https://doi.org/10.1007/s10654-007-9179-1

17. Pintar KDM, Pollari F, Waltner-Toews D, Charron DF, Mcewen SA, Fazil A, et al. A modified case-control study of cryptosporidiosis (using non-Cryptosporidium-infected enteric cases as controls) in a community setting. Epidemiol Infect [Internet]. 2009/06/16. 2009;137(12):1789–99. Available from: https://www.cambridge.org/core/article/modified-casecontrol-study-of-cryptosporidiosis-using-noncryptosporidiuminfected-enteric-cases-as-controls-in-a-community-setting/D41B4DC8FAEA4000BA4CD6A82D2F7934

18. Betancourt WQ, Rose JB. Drinking water treatment processes for removal of Cryptosporidium and Giardia. Vet Parasitol. 2004 Dec;126(1–2):219–34.

19. Hunter PR, Hughes S, Woodhouse S, Syed Q, Verlander NQ, Chalmers RM, et al. Sporadic cryptosporidiosis case-control study with genotyping. Emerg Infect Dis. 2004;10(7):1241–9.

20. Chalmers RM, Smith R, Elwin K, Clifton-Hadley FA, Giles M. Epidemiology of anthroponotic and zoonotic human cryptosporidiosis in England and Wales, 2004 to 2006. Epidemiol Infect. 2011 May;139(5):700–12.

21. Osborn A. Overseas residents in the UK and UK residents abroad: 2020 [Internet]. 2021. Available from: https://www.ons.gov.uk/peoplepopulationandcommunity/leisureandtourism/datasets/overseasresidentsintheukandukresidentsabroad

22. Statista. Holiday-related search terms with the highest year-over-year growth following the COVID-19 lockdown in the United Kingdom (UK) in July 2020 [Internet]. 2021 [cited 2021 Oct 8]. Available from: https://www.statista.com/statistics/1175666/growth-in-holiday-search-terms-uk/

23. Burnett H, Olsen JR, Nicholls N, Mitchell R. Change in time spent visiting and experiences of green space following restrictions on movement during the COVID-19 pandemic: a nationally representative cross-sectional study of UK adults. BMJ Open [Internet]. 2021 Mar 1;11(3):e044067. Available from: http://bmjopen.bmj.com/content/11/3/e044067.abstract

24. Natural England. People and Nature Survey: How has COVID-19 changed the way we engage with nature? [Internet]. 2022. Available from: https://naturalengland.blog.gov.uk/2022/05/18/people-and-nature-survey-how-has-covid-19-changed-the-way-we-engage-with-nature/

25. Office for National Statistics. How has lockdown changed our relationship with nature? [Internet]. London; 2021. Available from: https://www.ons.gov.uk/economy/environmentalaccounts/articles/howhaslockdownchangedourrelationshipwithnature/2021-04-26

26. Alzyood M, Jackson D, Aveyard H, Brooke J. COVID-19 reinforces the importance of handwashing. Vol. 29, Journal of clinical nursing. 2020. p. 2760–1.

27. Barrett C, Cheung KL. Knowledge, socio-cognitive perceptions and the practice of hand hygiene and social distancing during the COVID-19 pandemic: a cross-sectional study of UK university students. BMC Public Health [Internet]. 2021;21(1):426. Available from: https://doi.org/10.1186/s12889-021-10461-0

28. Love NK, Elliot AJ, Chalmers RM, Douglas A, Gharbia S, McCormick J, et al. Impact of the COVID-19 pandemic on gastrointestinal infection trends in England, February–July 2020. BMJ Open [Internet]. 2022 Mar 1;12(3):e050469. Available from: http://bmjopen.bmj.com/content/12/3/e050469.abstract

29. Robinson G, Elwin K, Chalmers RM. Cryptosporidium Diagnostic Assays: Molecular Detection BT - Cryptosporidium: Methods and Protocols. In: Mead JR, Arrowood MJ, editors. New York, NY: Springer New York; 2020. p. 11–22. Available from: https://doi.org/10.1007/978-1-4939-9748-0_2

30. Office for National Statistics. Mapping income deprivation at a local authority level [Internet]. 2019. Available from: https://www.ons.gov.uk/peoplepopulationandcommunity/personalandhouseholdfinances/incomeandwealth/datasets/mappingincomedeprivationatalocalauthoritylevel

31. Office for National Statistics. Rural/urban classifications [Internet]. 2011. Available from: https://www.ons.gov.uk/methodology/geography/geographicalproducts/ruralurbanclassifications

32. Bhaskaran K, Gasparrini A, Hajat S, Smeeth L, Armstrong B. Time series regression studies in environmental epidemiology. Int J Epidemiol [Internet]. 2013 Aug 1;42(4):1187–95. Available from: https://doi.org/10.1093/ije/dyt092

33. Kim H, Lee J-T, Fong KC, Bell ML. Alternative adjustment for seasonality and long-term time-trend in time-series analysis for long-term environmental exposures and disease counts. BMC Med Res Methodol [Internet]. 2021;21(1):2. Available from: https://doi.org/10.1186/s12874-020-01199-1

34. You C, Lin DKJ, Young SS. Time series smoother for effect detection. PLoS One. 2018;13(4):e0195360.

35. Cavanaugh JE, Neath AA. The Akaike information criterion: Background, derivation, properties, application, interpretation, and refinements. WIREs Comput Stat [Internet]. 2019 May 1;11(3):e1460. Available from: https://doi.org/10.1002/wics.1460

36. Chalmers RM, Elwin K, Thomas AL, Guy EC, Mason B. Long-term Cryptosporidium typing reveals the aetiology and species-specific epidemiology of human cryptosporidiosis in England and Wales, 2000 to 2003. Eurosurveillance. 2009;14(2):2.

37. Love NK, Elliot AJ, Chalmers RM, Douglas A, Gharbia S, McCormick J, et al. The impact of the COVID-19 pandemic on gastrointestinal infection trends in England, February – July 2020. medRxiv [Internet]. 2021 Jan 1;2021.04.06.21254174. Available from: http://medrxiv.org/content/early/2021/04/08/2021.04.06.21254174.abstract

38. Mertens G, Gerritsen L, Duijndam S, Salemink E, Engelhard IM. Fear of the coronavirus (COVID-19): Predictors in an online study conducted in March 2020. J Anxiety Disord [Internet]. 2020;74:102258. Available from: https://www.sciencedirect.com/science/article/pii/S0887618520300724

39. Rodríguez-Hidalgo AJ PY, D DI and F. Fear of COVID-19, Stress, and Anxiety in University Undergraduate Students: A Predictive Model for Depression. Front Psychol [Internet]. 2020;11:1–9. Available from: https://doi.org/10.3389/fpsyg.2020.591797

40. Public Health England. Cryptosporidium data 2008 to 2017 [Internet]. London; 2019. Available from: https://www.gov.uk/government/publications/cryptosporidium-national-laboratory-data/cryptosporidium-data-2008-to-2017#cryptosporidium-data-2008-to-2017

41. Gopfert A, Chalmers RM, Whittingham S, Wilson L, van Hove M, Ferraro CF, et al. An outbreak of Cryptosporidium parvum linked to pasteurised milk from a vending machine in England – a descriptive study, March 2021. Epidemiol Infect. 2022;

42. Chalmers RM, McCarthy N, Barlow KL, Stiff R. An evaluation of health protection practices for the investigation and management of Cryptosporidium in England and Wales. J Public Heal. 2016 Mar 1;388(1):1291–301.

43. Health Protection Agency. Standard Gastrointestinal Disease Questionnaire [Internet]. 2004 p. 2–4. Available from: https://assets.publishing.service.gov.uk/government/uploads/system/uploads/attachment_data/file/327958/gast_StandardGastro.pdf

